# Genomic insights into the comorbidity between type 2 diabetes and schizophrenia

**DOI:** 10.1101/2023.10.16.23297073

**Authors:** Ana Luiza Arruda, Golam M. Khandaker, Andrew P. Morris, George Davey Smith, Laura M. Huckins, Eleftheria Zeggini

## Abstract

Multimorbidity represents an increasingly important public health challenge with far-reaching implications for health management and policy. Mental health and metabolic diseases have a well-established epidemiological association. In this study, we investigate the genetic intersection between type 2 diabetes and schizophrenia. We use Mendelian randomization to examine potential causal relationships between the two conditions and related endophenotypes. We report no compelling evidence that type 2 diabetes genetic liability potentially causally influences schizophrenia risk and *vice versa*. Our findings show that increased body mass index (BMI) has a protective effect against schizophrenia, in contrast to the well-known risk-increasing effect of BMI on type 2 diabetes risk. We identify evidence of colocalization of association signals for these two conditions at 11 genomic loci, six of which have opposing directions of effect for type 2 diabetes and schizophrenia. To elucidate these colocalizing signals, we integrate multi-omics data from bulk and single-cell gene expression studies, along with functional information. We identify high-confidence effector genes and find that they are enriched for homeostasis and lipid-related pathways. We also highlight drug repurposing opportunities including N-methyl-D-aspartate (NMDA) receptor antagonists. Our findings provide insights into shared biological mechanisms for type 2 diabetes and schizophrenia, highlighting common factors that influence the risk of the two conditions in opposite directions and shedding light on the complex nature of this comorbidity.

## Introduction

Multimorbidity, the coexistence of two or more chronic health conditions within an individual, has been of growing public health concern in recent years ^1^. The simultaneous presence of multiple medical conditions poses substantial challenges for healthcare systems worldwide, affecting disease management, patient outcomes, and healthcare costs. Multimorbidity is not simply the sum of individual diseases but rather represents a complex interplay of interacting factors, including genetic predisposition, environmental influences, and shared pathophysiological pathways. Understanding the underlying mechanisms of multimorbidity is essential for guiding effective treatment strategies for patient-centered care. Yet, most health-related and drug development research is focused on treating or preventing individual diseases ^2^. Treating each condition separately is inefficient and increases the patient’s treatment burden, possibly leading to adverse effects.

Individuals with mental health disorders are at higher risk of having multimorbid physical health conditions than those without psychiatric disorders, which contribute to lower life quality and premature death ^3,4^. Here, we study the comorbidity between type 2 diabetes and schizophrenia, two conditions that commonly co-occur and have been described to be genetically correlated ^5^. Dissecting the shared genetic etiology of these diseases can help identify risk variants and effector genes that could be used as biomarkers or druggable targets for their treatment and prevention.

Type 2 diabetes is characterized by persistent elevated glucose levels and insulin resistance, with typical onset of symptoms during middle adulthood. In 2021, over 536 million people were affected by type 2 diabetes worldwide^6^. The heritability of type 2 diabetes has been estimated as ∼50%^7^. Schizophrenia is a major psychiatric disorder typically characterized by problems with perception, cognitive function and behavior and presents with hallucinations, delusion, disorganized thinking and speech^8^. Unlike type 2 diabetes, schizophrenia affects young people with onset during late adolescence or early adulthood. The prevalence of schizophrenia is ∼1%^9^, and of schizophrenia and related psychotic disorders is ∼3%. Genetic epidemiological studies have shown that schizophrenia has an estimated general heritability of ∼80% ^8^.

Observational studies have yielded indications of an epidemiologically positive association between type 2 diabetes and schizophrenia^10^. An evaluation of multiple observational studies derived a pooled relative risk of type 2 diabetes in schizophrenia patients versus healthy controls of 1.82 (95% confidence interval [CI]=1.56-2.13) ^11^. Sociodemographic and lifestyle factors are also considered key in the observed association. In addition, treatment with antipsychotic medication is associated with greater weight gain risk^12^ and increased risk for diabetes among individuals with schizophrenia^13^. However, hyperinsulinemia and impaired glucose tolerance have also been observed in first-episode schizophrenia patients who are antipsychotic naïve compared to healthy controls matched for age, sex and body mass index (BMI) ^14,15^. Genetic studies point to a partial shared genetic aetiology between type 2 diabetes and schizophrenia ^16-18^. For instance, a polygenic risk score for schizophrenia onset has been found to be positively correlated with insulin resistance among first-episode, drug naïve patients with schizophrenia ^19^. Causal inference analysis using Mendelian randomization has not provided evidence of a causal relationship between type 2 diabetes and schizophrenia^20^. Elevated fasting insulin levels have been shown to have a causal effect on schizophrenia, suggesting that insulin might exert an impact on this condition via a direct modulation of brain function instead of impaired glucose metabolism ^20,21^.

Beyond the role of increased adiposity, which is, in part, influenced by some antipsychotic drugs, possible additional biological pathways underlying the comorbidity between type 2 diabetes and schizophrenia may include immune dysfunction, particularly autoimmunity and chronic low-grade systemic inflammation^21,22^. Emerging evidence suggests that this interplay may be partly mediated by insulin resistance. Insulin has an anti-inflammatory effect, with inflammatory markers experiencing elevation in contexts of insulin resistance and diabetes ^23,24^. Increased levels of inflammatory mediators IL-1β, IL-6, TNF-α and pro-inflammatory adiponectin have been observed in drug naïve, first episode schizophrenia patients with normal weight compared to overweight individuals without schizophrenia ^25^. These findings have been reinforced by Mendelian randomization analyses that corroborate potential causal connections between these inflammatory biomarkers and schizophrenia^26^.

Genome-wide correlation analyses show evidence of a weak negative genetic correlation between type 2 diabetes and schizophrenia ^18,27^. Due to the difference in age-of-onset between the two conditions, and the diabetogenic side-effects of antipsychotic drugs, it is difficult to ascertain from observational studies whether type 2 diabetes and schizophrenia share underlying biological mechanisms. In this study, we aim to disentangle the shared genetic aetiology of type 2 diabetes and schizophrenia by resolving colocalizing signals to provide greater insight into the well-known comorbidity.

## Methods

### Datasets

#### GWAS summary statistics

For type 2 diabetes, we used the latest published multi-ancestry GWAS summary statistics not adjusted for BMI from the Diabetes Meta-Analysis of Trans-Ethnic (DIAMANTE) consortium that encompass 180,834 cases and 1,159,055 controls ^28^. The distribution of effective sample sizes across ancestries included 51.1% European ancestry, 28.4% East Asian ancestry, 8.3% South Asian ancestry, 6.6% African ancestry including admixed African American populations and 5.6% Hispanic ancestry. In our analyses utilizing the multi-ancestral findings, we integrated the p-values emanating from the meta-analysis conducted via MR-MEGA ^29^ and the effect size estimation and standard error from a fixed-effects model. We set the threshold for genome-wide significance at p-value < 5x 10^−8^. For schizophrenia, we used the latest published multi-ancestry GWAS summary statistics comprising data from 74,776 cases and 101,023 controls ^30^. The distribution across ancestries indicated European ancestry at 74.3%, East Asian ancestry at 17.5%, African American ancestry at 5.7%, and Latino ancestry at 2.5%. Genome-wide significance was set at p-value < 5x 10^−8^.

In some analyses, we also used GWAS summary data from type 2 diabetes and schizophrenia related traits. For type 2 diabetes we used data from glycaemic traits and adiposity-related traits. We used the following glycaemic traits data from MAGIC including up to 281,416 individuals without diabetes: fasting glucose, fasting insulin, HbAc1 and 2h-glucose-post-challenge^31^. Only summary statistics adjusted for BMI were made publicly available. Multi-ancestry summary statistics from MAGIC did not include effect size estimates, so we restricted the analyses to European ancestry summary statistics only. Adiposity-related traits were defined as BMI, body fat percentage, whole body fat mass and waist-to-hip ratio ratio (WHR) unadjusted for BMI. For BMI (N=806,834) and WHR (N=697,734), we used the recent meta-analysis combining data from the GIANT consortium and the UK Biobank ^32^. The inverse rank normalized GWAS summary statistics for whole body fat mass (N=330,762) and body fat percentage (N=331,117) from the UK Biobank were taken from the Neale Lab website (http://www.nealelab.is/uk-biobank/).

For schizophrenia, we analyzed GWAS summary statistics for attention deficit/hyperactivity disorder (ADHD), autism, anorexia nervosa, bipolar disorder, major depressive disorder, panic disorder/anxiety and post-traumatic syndrome disorder (PTSD). Data for these psychiatric traits were downloaded from the Psychiatric Genomics Consortium (PGC). For the psychiatric traits, sample sizes, references, and ancestry information from the GWAS summary statistics are summarized in **Table 1**.

**Table 1:** Overview of GWAS summary statistics of psychiatric traits employed in this study.

| Trait | N cases | N controls | Reference | Ancestry |
| --- | --- | --- | --- | --- |
| ADHD | 20183 | 35191 | Demontis *et al.*<sup>33</sup> | European + Danish |
| Anorexia nervosa | 16992 | 55525 | Watson *et al.*<sup>34</sup> | European |
| Autism | 18381 | 27969 | Grove *et al.*<sup>35</sup> | Danish |
| Bipolar disorder | 41917 | 371549 | Mullins *et al.*<sup>36</sup> | European |
| Major depressive disorder | 170756 | 329443 | Howard *et al.*<sup>37</sup> | European |
| Panic disorder/anxiety | 2248 | 7992 | Forstner *et al.*<sup>38</sup> | European |
| PTSD | 32428 | 174227 | Nievergelt *et al.*<sup>39</sup> | Multi-ancestry |

#### Molecular quantitative trait loci summary statistics

We used various datasets of molecular quantitative trait loci (QTLs) from relevant tissues for each condition, namely brain (adult and fetal) for schizophrenia and adult brain, pancreatic islets, liver and subcutaneous/visceral adipose tissue for type 2 diabetes. We used bulk and single-cell type data from different molecular levels, including expression (eQTL), protein (pQTL), chromatin accessibility (caQTL), methylation (mQTL) and splicing (sQTL). For the eQTLs from the brain cortex, summary statistics were available for different ancestries separately. GTEx v8 expression and splicing QTL datasets consist of multi-ancestry samples, the majority (85.3%) of which are of European ancestry. All other data sets are from individuals of European ancestry only. A detailed list of the employed QTL datasets including sample size can be found in **Table 2**.

**Table 2:**
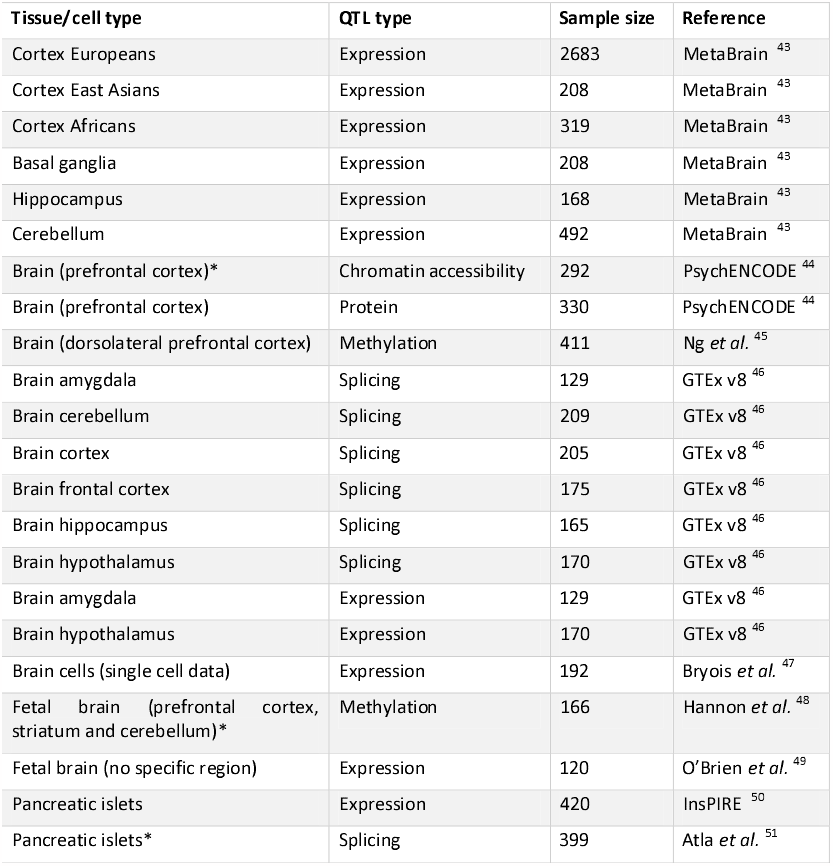
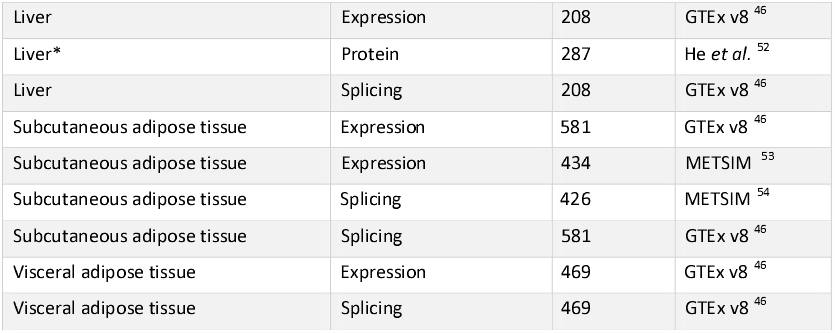
Overview of molecular quantitative trait loci (QTL) summary statistics employed in this work.

For the brain caQTL, fetal brain mQTL, pancreatic islets sQTL and liver pQTL, only genome-wide significant or nominally significant results were available. We performed a lift-down from GRCh38 to GRCh37 using the R package *CrossMap* (version 0.5.4) for all eQTL datasets from MetaBrain and the fetal brain eQTL dataset ^40^. We extracted regional expression and splicing QTL data from GTEx v8 by querying the eQTL catalogue’s RESTful application programming interface (API) v2 in R ^41^. Subsequently, leveraging the *liftOver* function from the R package *rtracklayer* (version 1.22.0) ^42^, we conducted a genomic mapping from GRCh38 to GRCh37. For GTEX v8, splicing QTLs were defined as leafcutter splice junction QTLs. Leafcutter quantifies RNA splicing variation using short-read RNA-seq data.

### Quantification and statistical analysis

#### Genetic overlap between type 2 diabetes and schizophrenia

We quantified genetic correlation between type 2 diabetes and schizophrenia by conducting a linkage disequilibrium (LD) score regression analysis using the LDSC software (v1.0.1) with –rg flag ^55^. We used the multi-ancestry GWAS summary statistics from the meta-analyses for type 2 diabetes and schizophrenia to enhance our discovery power. Since the European ancestry proportion is the largest one, we used the pre-computed LD scores from the 1000 Genomes European ancestry haplotypes ^56^. A sensitivity analysis was conducted using exclusively European subset summary statistics. The complete results for the primary and the sensitivity analyses can be found in **Table S1**.

#### Causal inference analysis between schizophrenia and type 2 diabetes

To assess whether schizophrenia and type 2 diabetes have a causal relationship, we performed bi-directional two-sample Mendelian randomization analyses using the multi-ancestry GWAS summary statistics to enhance predictive power ^57^ (**Table S2A**). We used the *TwoSampleMR* R package (version 0.5.7), which is curated by MR-Base ^58^. We selected independent genome-wide significant (p-value < 5x 10^−8^) variants as instrumental variables (IVs). Independence was defined as LD-based clumped variants with a strict LD threshold of R^2^ = 0.001 over a 10Mb window on either side of the index variant. Since the largest proportion of the samples in both type 2 diabetes and schizophrenia GWAS summary statistics are from European ancestry, we used the European reference LD panel from 1000 Genomes. As a sensitivity analysis for the estimated direction of effect, we applied Steiger-filtering to ensure that the IVs were more strongly associated with the exposure than the outcome (**Table S2B**). Additionally, we performed sensitivity analyses on the European ancestry subset only to compare the direction of effect (**Table S2C**).

To assess whether the IVs were strongly associated with the exposure, we calculated the F-statistic for each IV and selected only those that were larger than ten for the Mendelian randomization analyses to minimise bias due to weak IVs. F-statistic was estimated from summary level data as (*beta*^2^*/se*^2^), where beta is the effect size and se is the standard error ^57^. After the removal of weak IVs, we calculated the overall F-statistic as *mean*(*beta*^2^ */se*^2^). We applied the inverse variance weighted (IVW) method, which performs a random-effects meta-analysis of the Wald ratios for each SNP. As a first sensitivity analysis, we applied the weighted median (WM) and the MR-Egger regression methods to ensure consistency of the effect size direction. The intercept of MR-Egger regression was used to assess horizontal pleiotropy. Finally, we tested for heterogeneity using the Q-statistic, which was calculated using the *mr_heterogeneity* function from the *TwoSampleMR* R package. To account for multiple testing, we corrected the p-values separately for each Mendelian randomization method employed by using the Bonferroni method. We extended the causal inference analyses to other psychiatric traits (**Table 1**) and endophenotypes related type 2 diabetes including adiposity-related traits.

#### Causal inference using childhood and adulthood body mass index as exposures

We performed causal inference analysis using univariate and multivariate two-sample Mendelian randomization with childhood and adulthood BMI as exposures. For childhood BMI, we used the IVs derived from a genetically predicted early life body size GWAS calculated based on recall data, from the UK Biobank^59^. Firstly, we estimated the total effect of each life-stage BMI value by conducting univariate Mendelian randomization analyses using each genetically predicted BMI value as a separate exposure and type 2 diabetes and schizophrenia as outcomes (**Table S3**). For this analysis, we applied two-sample Mendelian randomization with the IVW, WM and MR-Egger regression methods using the *TwoSampleMR* R package, as described above^58^.

Subsequently, we performed a multivariate Mendelian randomization analysis to estimate the direct effects of each life-stage BMI value using both childhood and adulthood BMI simultaneously as exposures (**Table S4**). We used the LD independent IVs from the univariate analyses. We applied the IVW method from the *MVMR* R package (version 0.4)^60^. To assess the effect of heterogeneity due to pleiotropy on the causal estimates, the *MVMR* R package calculates an adapted heterogeneity Q-statistic and estimates the causal effect of each exposure on the outcome accounting for heterogeneity, pleiotropy, and weak instrument bias. For both univariate and multivariate analyses we conducted sensitivity analyses for the direction of effect using only the European subset of the type 2 diabetes and schizophrenia GWAS (**Table S3B** and **Table S4B**).

#### Genetic colocalization analysis

For both type 2 diabetes and schizophrenia, we defined genomic regions spanning 2Mb windows centered on independent genome-wide significant lead SNPs from the individual GWAS summary statistics. Within each identified region, we performed statistical colocalization analysis between type 2 diabetes and schizophrenia using the estimated regression coefficients (effect sizes) and standard errors (**Table S5**). We used the *coloc*.*abf* function from the *coloc* R package (version 3.2.1) for the analyses ^61^. This function calculates the posterior probability for a set of five association hypotheses under the assumption of a single causal variant per trait in the region. Our investigative emphasis lay primarily upon the broader genetic landscape encompassing these regions, rather than a focused endeavour to identify a precise causal variant. Thus, the single-variant assumption of *coloc*.*abf* was not an issue here. The hypotheses are summarized below:

***H0:*** no trait has a genetic association in the region

***H1:*** trait 1 has a genetic association in the region

***H2:*** trait 2 has a genetic association in the region

***H3:*** both traits have a genetic association in the region, but with different causal variants

***H4:*** both traits share a genetic association (single causal variant) in the region

We used the default prior probabilities of the *coloc* R package for our analyses. We considered evidence for colocalization if the posterior probability of H4 (PP4) > 0.8. In addition to the posterior probability, we calculated a 95% credible set for the causal variant by taking the cumulative sum of the variants’ posterior probabilities to be causal, conditional on H4 being true. LD between the estimated lead causal variant and the other variants in the region was calculated using PLINK (version 2.0 alpha) ^62^ based on the UK Biobank data ^32^ and was used for visualizing the results in regional association plots. In regions where we find evidence of colocalization (PP4>0.8), we ran sensitivity analyses using GWAS summary statistics for type 2 diabetes and schizophrenia derived from samples of European ancestry only.

#### Knockout mouse phenotypes

We performed a schematic search for each gene located within the vicinity (2 Mb window) of the genomic loci that colocalized between type 2 diabetes and schizophrenia to screen for knockout mice showing phenotypes related to each of the conditions. The databases used in this scope were the International Mouse Phenotyping Consortium (IMPC) (https://www.mousephenotype.org/), Mouse Genome Informatics (MGI) (http://www.informatics.jax.org/) and Rat Genome Database (RGD) (https://rgd.mcw.edu/) databases. For IMPC and RGD we extracted the knockout mice phenotypes for each analysed gene using the programmatic data access via their API. For MGI, we used the MGI batch query.

For type 2 diabetes, we looked for insulin and diabetes-related phenotypes that included the following terms: insulin, glucose, diabetes, hyperglycaemia, pancreas, pancreatic, obesity, BMI, body weight, body mass, body fat, beta cell, and glucosuria. For schizophrenia, we included phenotypes related to brain and craniofacial morphology, behaviour and psychiatric traits. We based our inclusion criteria on the Five Factor Model used to diagnose schizophrenia in humans, which does not include motoric phenotypes ^63^. As a sensitivity analysis we extended the list of phenotypes related to schizophrenia to motoric phenotypes. In addition to phenotypes covered by the Five Factor Model, we included craniofacial morphology and eye movement phenotypes in line with the literature and previous schizophrenia-related work ^64-66^. A full list of the included phenotypes can be found in **Table S9A** and **Table S9B**.

#### Rare and syndromic human diseases

For every gene located within the vicinity of genomic regions that show evidence of genetic colocalization between type 2 diabetes and schizophrenia, we extracted data from the Online Mendelian Inheritance in Man (OMIM) (https://omim.org/) via its API. We looked up whether any rare and syndromic diseases linked to those genes showed any phenotype related to type 2 diabetes or schizophrenia. We defined association with schizophrenia if the disease showed any neurological phenotype. The full list of rare and syndromic human diseases associated with type 2 diabetes and schizophrenia can be found in **Table S9A** and **Table S9B** respectively.

#### Differential gene expression

For the genes in the regions of colocalization between type 2 diabetes and schizophrenia, we conducted a lookup on publicly available summary statistics of differential expression RNA-seq datasets related to the studied conditions. For type 2 diabetes, we used RNA-seq data from surgical pancreatic tissue samples from 57 metabolically phenotyped pancreatectomized patients, from which 39 were previously diagnosed with type 2 diabetes and 18 were non-diabetes patients ^67^. The differential expression analysis was based on a linear model with age, sex, and BMI as covariates. We defined genes as differentially expressed if they changed more than 1.5-fold in either direction and had an adjusted p-value < 0.05, as in the original publication. For schizophrenia, we retrieved differential gene expression data based on RNA-seq and genotype data of post-mortem brains (frontal and temporal cortex) from PsychENCODE that includes 559 schizophrenia patients and 936 controls ^68^. Differential expression was assessed using a linear mixed effects model accounting for known biological, technical and surrogate variables as fixed effects as well as subject-level technical replicates as random effects. Genes were defined as differentially expressed at a false discovery rate (FDR) < 0.05.

#### Multi-trait colocalization analysis with QTL data

Within the genomic loci exhibiting evidence of colocalization between type 2 diabetes and schizophrenia, we performed multi-trait colocalization analyses between type 2 diabetes GWAS summary statistics, schizophrenia GWAS summary statistics and each molecular QTL summary statistics from disease-relevant tissues or cell types summarized in **Table 2**. The analyses were performed within a 2Mb window around the lead variant of the 95% credible set from the colocalization analysis between type 2 diabetes and schizophrenia.

To perform the multi-trait colocalization analyses, we used the R package *HyPrColoc* (version 1.0) ^69^. Similarly to the *coloc*.*abf* function from the *coloc* R package, *HyPrColoc* estimates posterior probabilities for the above-mentioned colocalization hypotheses and identifies a putative shared causal variant. To assess whether all traits colocalize together without grouping them, we switched off the Bayesian divisive clustering algorithm (*bb*.*alg* = FALSE). As instructed by the developers, we assumed no sample overlap between all the input summary statistics. Evidence of colocalization was defined for a PP4 > 0.8. LD between the lead causal variant and the other variants in the genomic locus was calculated using PLINK (version 2.0 alpha) ^62^ based on the UK Biobank data ^32^.

Since we know a priori that schizophrenia and type 2 diabetes signals colocalize in the analysed genomic loci, we adapted the prior probabilities of the *HyPrColoc* algorithm accordingly. For the first parameter, *prior*.*1*, which denotes the probability of a variant being associated with a single trait, we kept the default value of 10^−4^ since most regions that colocalize between type 2 diabetes and schizophrenia are genome-wide significant (p-value < 5x 10^−8^) for only one of the conditions. We slightly increased the conditional colocalization prior parameter, *prior*.*c*, from the default of 0.02 to 0.05 due to the evidence of colocalization between type 2 diabetes and schizophrenia in the region. This parameter represents the prior probability that a variant is associated with an additional trait given that it is associated with one trait.

As for the colocalization between type 2 diabetes and schizophrenia, we calculated a 95% credible set for the causal variant for each colocalized genomic locus by taking the cumulative sum of the variants’ posterior probabilities to be causal conditional on H4 being true. We considered relevant evidence of colocalization between distinct QTL datasets and both type 2 diabetes and schizophrenia if the 95% credible set of the *HyPrColoc* colocalization overlapped at least by one variant with the 95% credible set of the type 2 diabetes-schizophrenia colocalization analysis.

#### Scoring of potential effector genes

Firstly, we scored all genes within the vicinity (2Mb window centered on the lead causal variant) of genome loci that colocalize (PP4 > 0.8) between type 2 diabetes and schizophrenia using four biological lines of evidence based on multi-omics and functional biological data:

1. Colocalization with at least one molecular QTL from disease-relevant tissues/cell types
2. Differential gene expression from pancreatic islets or brain
3. Knockout mice with phenotypes related to the type 2 diabetes or schizophrenia
4. Rare and syndromic human diseases with phenotypes related to the type 2 diabetes or schizophrenia

For colocalization with methylation QTLs, we used the *getMappedEntrezIDs* function from the *missMethyl* R package (version 1.32.1) using annotation data from the Illumina’s HumanMethylation450 platform to map CpG sites to genes ^70^. We excluded the following genes from the analysis due to inconsistent gene identifiers: *SNORA40, snoU13* and *Y_RNA*.

We constructed one score for each condition. Each score was extended by information on established effector genes for the corresponding condition. For type 2 diabetes, we retrieved a list of 135 high-confidence effector genes from the curated T2D Effector Prediction Summary from the Type 2 Diabetes Knowledge Portal (https://t2d.hugeamp.org/method.html?trait=t2d&dataset=egls) that scored at least 3. For schizophrenia, we used a list of 321 established effector genes from PsychENCODE that were supported by more than two evidence sources ^44^. Since our analysis overlaps with criteria used to define a gene as high-confidence for the individual conditions, we followed an orthogonal approach to incorporate this information: if a gene is high-confidence for a condition, but scored zero in our analysis, we updated the respective condition score to one.

As an additional line of evidence, we looked up whether any variant in the 95% credible set from the colocalization analysis between type 2 diabetes and schizophrenia was a missense variant for any of the analyzed genes on Ensembl GRCh37 release 110 ^71^. The total score was defined as the sum of the type 2 diabetes, schizophrenia and missense score. If the missense variant score was the only line of evidence for a gene, the total score was kept at zero since this information is not directly related to the studied comorbidity.

We scored the genes based on the outlined six biological lines of evidence that were integrated in an orthogonal manner (**Table S6**). Genes that scored at least one point in the type 2 diabetes score andone point in the schizophrenia score were defined as potential effector genes for the comorbidity. From those genes, the ones that scored at least 3 points in the total score were defined as likely effector genes. Finally, a subset of those that scored at least 4 were defined as high-confidence effector genes.

#### Causal inference analysis using multi-omics data

To query whether the high-confidence genes have a causal effect on type 2 diabetes or schizophrenia, we performed two-sample Mendelian randomization analyses between the expression QTL summary statistics and each disease GWAS summary statistics (**Table S7A**). We conducted Mendelian randomization analyses with different molecular QTL of tissues or cell types that show evidence of colocalization with both conditions simultaneously. We used the same software and sensitivity analyses described above for the causal inference analysis between type 2 diabetes and schizophrenia. If only one IV was available after clumping, harmonizing the data and filtering weak IVs through the F-statistics, we employed the Wald ratio method. We were only able to perform MR-Egger regression in instances where there were more than three independent IVs. If the IVs from the QTL datasets were absent in the disease GWAS, we ran the Mendelian randomization analysis with a LD proxy variant calculated on the European ancestry 100 genome reference panel instead. To search for LD proxies, we used the R package *LDlinkR* that queries the web-based LDlink tool and based the calculations in the European ancestry 100 genome reference panel ^72^. The adjusted significance threshold for the eQTL data from adipose tissue from METSIM was set to p-value < 5x 10^−^, as defined in the original publication ^53^. We performed sensitivity analyses within the European ancestry subset only to compare the direction of effect (**Table S7B**).

#### Pathway analysis

On the derived likely effector genes set for the comorbidity between type 2 diabetes and schizophrenia, we performed gene set enrichment analysis using the human resources and the enrichment software from the ConsensusPathDB (http://cpdb.molgen.mpg.de/) ^73^ (**Table S8**). We did not perform the analysis on the high-confidence effector genes due to the small number of genes on this set. For the enrichment analysis, we used the Gene Ontology human networks including following subcategories up to level 5: molecular function, biological processes, and cellular component. Finally, we required a minimum overlap of 2 genes for enrichment and set the significance threshold at FDR < 0.05.

#### Druggable genome

We queried the druggability status of the likely effector genes for the type 2 diabetes and schizophrenia comorbidity using two databases. Firstly, we used the Druggable Genome database that consists of 4479 genes classified into three tiers based on their progress in the drug development pipeline ^74^. Tier 1 consists of 1427 genes that are targets or clinical-phase drug candidates of already approved small molecules and biotherapeutic drugs. Tier 2 includes 682 genes that encode targets with known bioactive drug-like small-molecule binding partners and genes with ≥ 50% identity (over ≥75% of the sequence) with approved drug targets. Tier 3 comprise 2370 genes that encode secreted or extracellular proteins, proteins with more distant similarity to approved drug targets, and members of key druggable gene families that were not included in Tier 1 or 2. Tier 3 was further subdivided to prioritize genes in proximity (+-50 kbp) to a GWAS risk variant based on data from the GWAS catalog and had an extracellular location (Tier 3A). Tier 3B consists of the remaining genes.

For likely effector genes included in Tier 1 from the Druggable Genome, we further examined the approved or clinical trial drugs using the DrugBank database (https://www.drugbank.com, accessed on the 9^th^ of August 2023) and the Open Targets platform ^75^.

## Results

### Insights into shared genetics between type 2 diabetes and schizophrenia

We assessed the genetic correlation between type 2 diabetes (*N*_*cases*_ = 180,834, *N*_*controls*_ = 1,159,055) and schizophrenia (*N*_*cases*_ = 74,776, *N*_*controls*_ = 101,023) on a genome-wide scale using multi-ancestry data from the largest published GWAS summary statistics for the individual conditions. Using the same data, we assessed the causal relationship via Mendelian randomization analyses between type 2 diabetes and schizophrenia on a genome-wide scale. We find nominal evidence for a negative genetic correlation (r_g_ = −0.04, standard error = 0.02, p-value = 0.023) (**Table S1**) and no evidence of a causal relationship between type 2 diabetes and schizophrenia (**Table S2A**). As a result of insufficient monitoring of patients, potential inability, or unwillingness to seek help, and poor integration of health care services, comorbidities are often underdiagnosed among schizophrenia patients^76^. Hence, we expanded the Mendelian randomization analysis to endophenotypes of type 2 diabetes including adiposity-related traits. We find evidence that increased BMI is protective against schizophrenia (BMI ⟶ schizophrenia: odds ratio (OR)=0.81, 95% CI=(0.74,0.89), adjusted p-value (p.adj)=2.26 × 10^−4^) and schizophrenia liability is protective against increased BMI (schizophrenia ⟶ BMI: OR=0.98, 95% CI=(0.96,0.99), p.adj=0.03). For the BMI to schizophrenia analysis, the effect remained protective in the European-only sensitivity analysis (**Table S2C**), but it changed direction after Steiger-filtering the data (**Table S2B**). For the schizophrenia to BMI analysis, the protective direction of effect remained consistent in the Steiger-filtered analysis (**Table S2B**) and in the analysis using only the European subset of the schizophrenia GWAS summary statistics (**Table S2C**).

To further dissect the effect of BMI on type 2 diabetes and schizophrenia, we performed causal inference analyses using Mendelian randomization in a univariate and multivariate setting with childhood and adulthood BMI as exposures (**Figure 1**Error! Reference source not found.). For type 2 diabetes, we find that both childhood and adulthood BMI are causal for the disease when studied individually (childhood BMI: OR=2.59, 95% CI=(2.23,3.01), p.adj=1.39 × 10^−^ ; adulthood BMI: OR=3.58, 95% CI=(3.09,4.16), p.adj=4.51 × 10^−^) (**Table S3**). We replicated previous results^77^ and show that the effect of childhood BMI is completely attenuated in the multivariate setting (childhood BMI: OR=1.19, 95% CI=(0.95,1.5), p.adj=0.18; adulthood BMI: OR=3.27, 95% CI=(2.68,3.99), p.adj=1.26 × 10^−30^) (**Table S4**). For schizophrenia, we find that only adulthood BMI has a significant protective effect against the condition in both the univariate (childhood BMI: OR=0.88, 95% CI=(0.72,1.06), p.adj=0.17; adulthood BMI: OR=0.75, 95% CI=(0.64,0.87), p.adj=1.99 × 10^−4^) (**Table S3**) and multivariate setting (childhood BMI: OR=1.04, 95% CI=(0.8,1.34), p.adj=0.79; adulthood BMI: OR=0.77, 95% CI=(0.61,0.96), p.adj=0.046) (**Table S4**).

**Figure 1:**
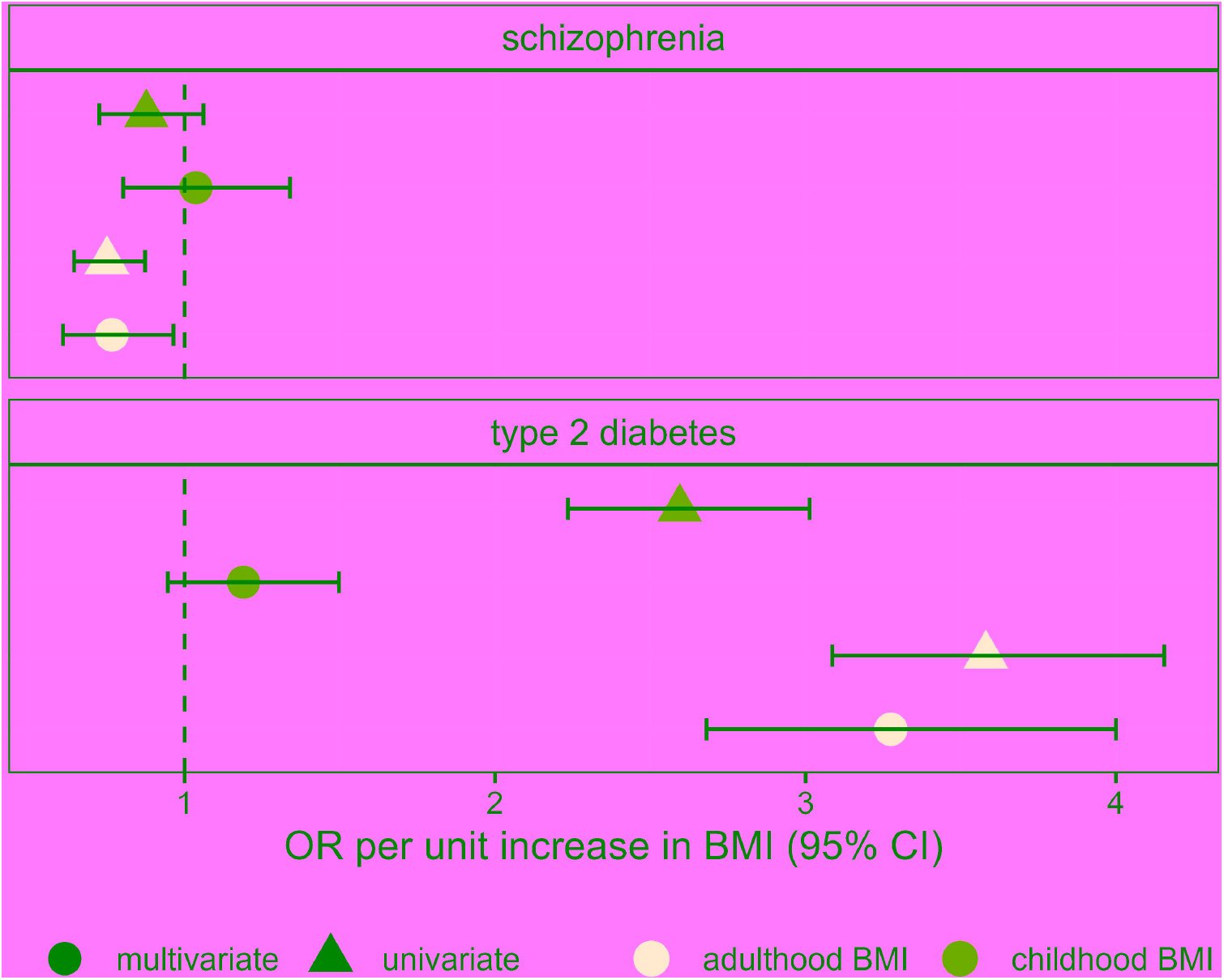
Mendelian randomization results of childhood versus adulthood BMI analysis. Forest plot depicting the direct and indirect effects for genetically predicted childhood and adulthood BMI on type 2 diabetes and schizophrenia. The effect is shown in odds ratio (OR) per unit increase in the BMI category, namely childhood and adulthood BMI. The results of the univariate analysis are represented by triangles and the multivariate results are represented by circles.

To identify shared genetic signals, we performed Bayesian colocalization analysis on genome-wide significantly associated type 2 diabetes and schizophrenia risk loci (p-value < 5x 10^−8^) using the GWAS summary statistics for each condition. We find evidence of colocalization (posterior probability of a shared causal variant (PP4) > 0.8) in 11 genomic loci (**Table S5**). Six of these genomic loci have opposing directions of effect for type 2 diabetes and schizophrenia. For two loci, the 95% credible set for the causal variant of the colocalization analysis consists of only one genetic variant, narrowing down the common genetic risk to a single variant.

To resolve colocalizing signals, we incorporated multi-omics and functional biology information to score the 444 genes located within the 11 genomic loci that colocalized between type 2 diabetes and schizophrenia (**Figure 2**). We performed multi-trait colocalization analyses with bulk and single-cell molecular QTL from disease-relevant tissues and cell-types, namely several brain regions for schizophrenia, and pancreatic islets, liver, and subcutaneous adipose tissue for type 2 diabetes, respectively (**Figure 2**). Our analyses included data from expression, protein, splicing and chromatin accessibility QTLs. In addition, we assessed whether these genes were differentially expressed in pancreatic islets or brain. We further conducted an extensive phenotypic search on knockout mice and rare and syndromic human diseases databases. Finally, we annotated missense variants in the 95% credible sets from the colocalization. 37 genes showed at least one line of evidence linking them to type 2 diabetes and at least one linking them to schizophrenia and were defined as potential effector genes for the comorbidity. Of these, we defined 15 likely effector genes that had a score of at least three, and three high-confidence effector genes that displayed at least four different lines of evidence, namely *EGR2, LAMA4* and *NUS1* (**Table S6**). None of the three high-confidence effector genes has been previously defined as high-confidence for either type 2 diabetes or schizophrenia.

**Figure 2:**
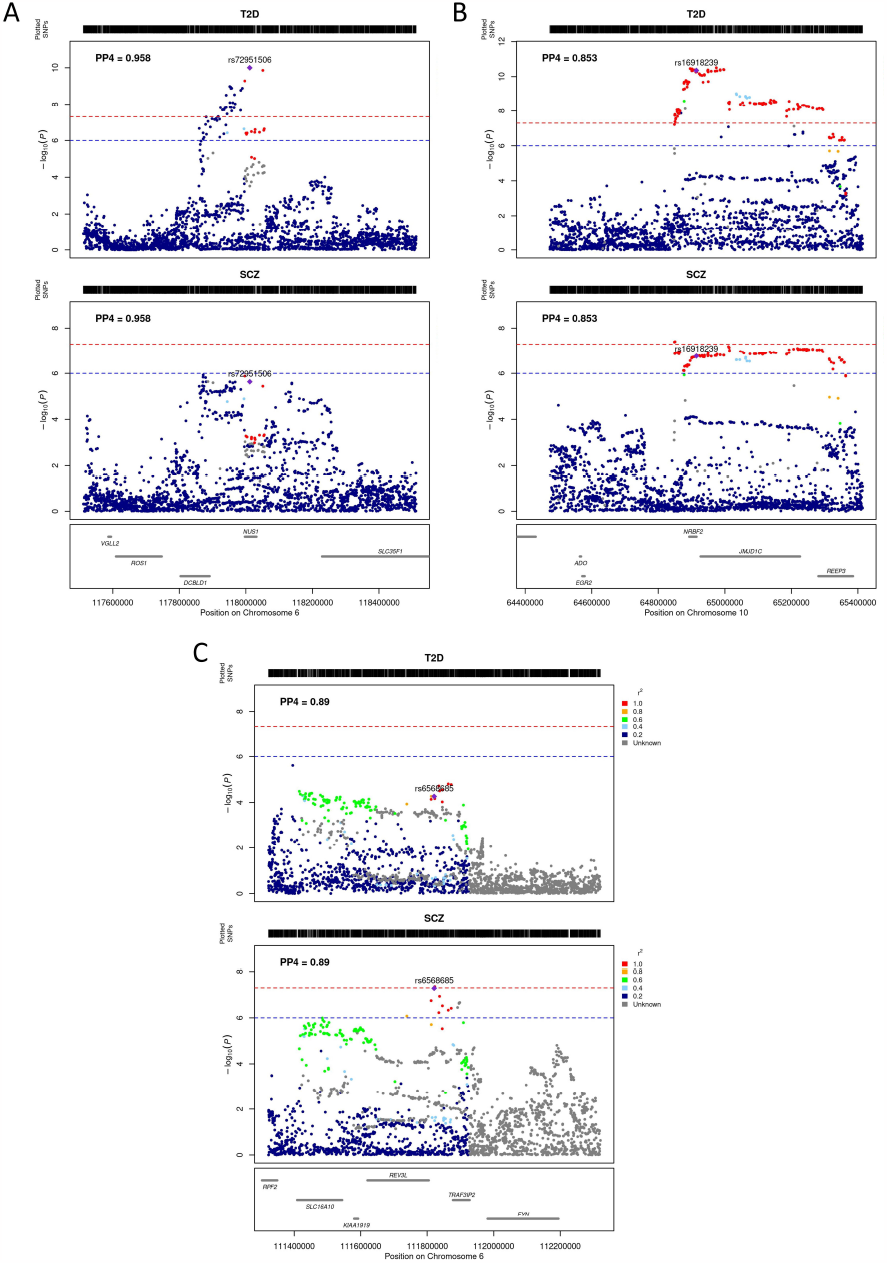
Study design for scoring genes around colocalized regions. Created with BioRender.com.

### Insights gained from high-confidence effector genes

#### NUS1

*NUS1* (nuclear undecaprenyl pyrophosphate synthase 1 homolog) is the highest-scoring effector gene. This genomic locus colocalizes between type 2 diabetes and schizophrenia with a PP4 of 0.96 and includes six variants in the 95% credible set for the shared causal variant from the colocalization (**Figure 3A**). All six variants have opposing risk-increasing alleles for type 2 diabetes and schizophrenia. The lead causal variant, rs72951506, is located in the intron of *NUS1* and reaches nominal significance for schizophrenia (p-value = 2.3 × 10^−^) and genome-wide significance for type 2 diabetes (p-value = 9.93 × 10^−11^). Additionally, type 2 diabetes and schizophrenia colocalize with genetic variants associated with the expression of *NUS1* in the cerebellum (PP4 = 0.94), cortex (PP4 = 0.89) and pancreatic islets (PP4 = 0.96). The variants in the 95% credible set from the colocalization are associated with increased expression of *NUS1* in these tissues, increased risk of schizophrenia and decreased risk of type 2 diabetes (**Table 3**).

**Table 3:**
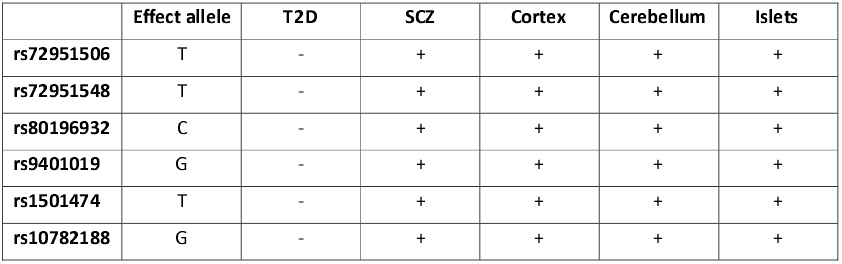
Direction of effect of the six variants included in the 95% credible set from the colocalization between type 2 diabetes and schizophrenia. The table depicts the direction of effect of six variants for type 2 diabetes (T2D), schizophrenia (SCZ), gene expression in the cortex, cerebellum and pancreatic islets.

**Figure 3:**
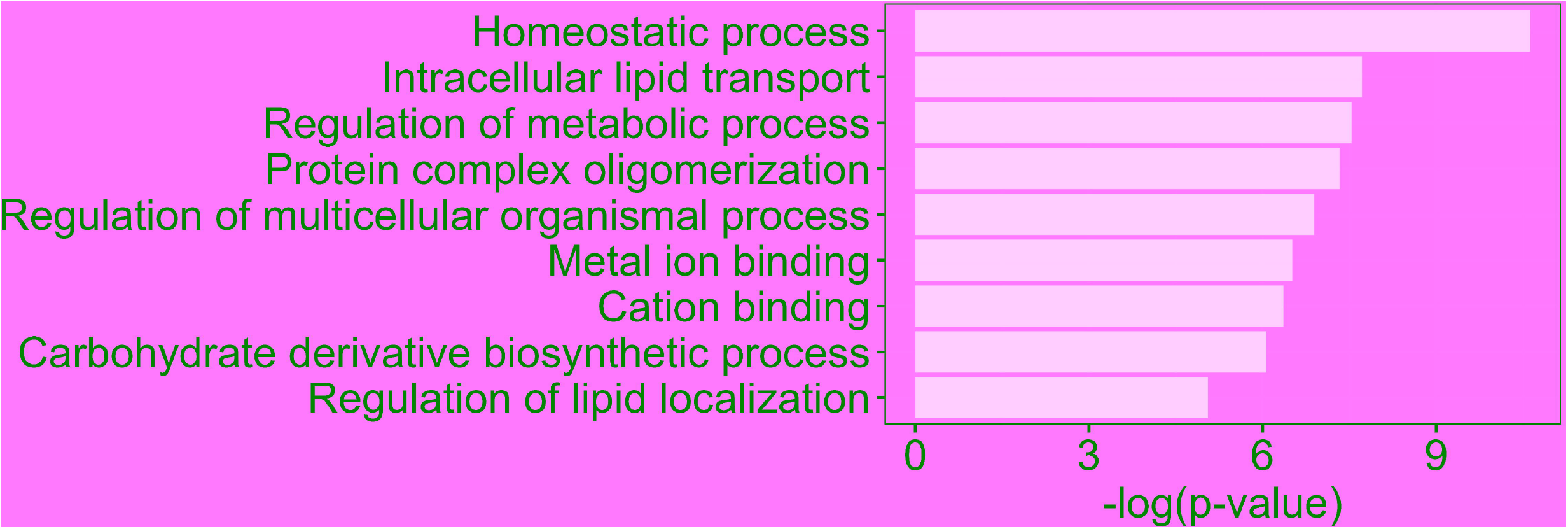
Regional plots of colocalized regions that yielded a high-confidence effector gene. A) NUS1 region, B) EGR2 region, C) LAMA4 region.

*NUS1* encodes a type I single transmembrane domain receptor, which is a subunit of cis-prenyltransferase, and serves as a specific receptor for the neural and cardiovascular regulator Nogo-B. ^78^. It is involved the regulation of intracellular LDL-derived cholesterol trafficking ^79^. Variants in the *NUS1* gene are associated with type 2 diabetes, epilepsy and intellectual disability. *NUS1* has not been previously defined as a high-confidence gene for type 2 diabetes nor schizophrenia and is not a target of any approved drug.

*NUS1* shows significantly higher expression in brains from schizophrenia patients compared to controls ^68^. *Nus1* knockout mice show increased circulating triglyceride levels, abnormal brain vasculature morphology and abnormal lipid homeostasis. Increased levels of circulating triglycerides have been shown to be associated with increased risk of type 2 diabetes in humans ^80^. Two rare and syndromic human diseases are associated with mutations in *NUS1*. The first, congenital disorder of glycosylation type 1aa, is a severe neurodevelopmental disorder with symptoms including seizures, developmental delay and hypotonia ^78^caused by a homozygous mutation in the *NUS1* gene. The mutation results in an Arg290His substitution at a residue in the highly conserved C-terminal domain. The second syndromic disease associated with *NUS1* is intellectual development disorder-55 with seizures. It is caused by de-novo heterozygous mutations in the *NUS1* gene ^81^. The mutations leading to both disorders result in autosomal-dominant loss-of-function variants in *NUS1* ^82,83^, which contradicts the findings from the differential gene expression analysis in brains of schizophrenia patients compared to controls. Fibroblasts with silenced *NUS1* show increased accumulation of free cholesterol ^78^.

We performed Mendelian randomization between the expression of high-confidence effector genes and type 2 diabetes or schizophrenia (**Table S7A**). Our analysis results suggest an opposing direction of causal effect of *NUS1* expression on type 2 diabetes and schizophrenia risk. Increased expression of *NUS1* in the brain and in pancreatic islets has a causal effect on schizophrenia (cortex: OR=1.22, 95% CI=(1.09,1.37), p.adj=0.002; cerebellum: OR=1.05, 95% CI=(1.02,1.09), p.adj=0.002; islets: OR=1.21, 95% CI=(1.12,1.3), p.adj < 9 × 10^−^). Differential expression analysis in brain supports this evidence, as *NUS1* is over-expressed in schizophrenia patients. Increased expression of *NUS1* in brain and pancreatic islets is protective against type 2 diabetes (cortex: OR=0.92, 95% CI=(0.85,0.99), p.adj=0.06; cerebellum: OR=0.94, 95% CI=(0.94,0.97), p.adj < 7 × 10^−^ ; islets: OR=0.86, 95% CI=(0.82,0.9), p.adj = 6 × 10^−9^).

#### EGR2

A genomic locus on chromosome 10 that encompasses a high-confidence gene colocalizes between type 2 diabetes and schizophrenia with a PP4 of 0.84 (**Figure 3B**). This region shows PP4=0.082 and PP3=0.82 in the European ancestry-only sensitivity analysis. The 95% credible set from the colocalization analysis contains 56 variants. Synthesis of the orthogonal lines of evidence from functional genomics data point to *EGR2* (early growth response-2) as the high-confidence effector gene for this region. For all shared causal candidate variants, the risk-increasing alleles for type 2 diabetes and schizophrenia are opposite.

*Egr2* knockout mice show decreased body weight, weight loss, decreased nerve conduction velocity, decreased neuron number, delayed eye opening, and abnormal neuron physiology and differentiation. In humans, these phenotypes are associated with a lower risk of type 2 diabetes and a higher risk of schizophrenia. *EGR2* shows significantly lower expression in the brain of schizophrenia patients compared to healthy controls ^68^.

Mutations in the *EGR2* gene are associated with three rare and syndromic human diseases: Charcot-Marie-tooth disease type 1D, a sensorineural peripheral polyneuropathy that affects both motor and sensory nerve function; Dejerine-Sottas syndrome, a demyelinating peripheral neuropathy with onset in infancy that results in delayed motor development; and Congenital hypomyelinating neuropathy, which is characterized clinically by onset of hypotonia at birth, areflexia, distal muscle weakness, and very slow nerve conduction velocities. In all these diseases, the mutation of *EGR2* leads to a decrease or a complete loss of gene function.

*EGR2* encodes a transcription factor that is a prime regulator of Schwann cell myelination ^84^. It is involved in the development of the jaw opener musculature by playing a role in its innervation through trigeminal motor neurons ^85^. *EGR2* has been reported to also play a role in hindbrain segmentation and development ^84^ and in adipogenesis, possibly by regulating the expression of CEBPB ^86^. Variants in *EGR2* gene are associated with serum triglycerides and cholesterol levels, temperament and adventureness.

Mendelian randomization analysis shows evidence of causality between increased expression of *EGR2* in the cortex and schizophrenia (OR=1.21, 95% CI=(1.09,1.35), p.adj=0.002) (**Table S7A**). There was no evidence of a causal effect of *EGR2* levels in disease-relevant tissues on type 2 diabetes.

#### LAMA4

We identify *LAMA4* (laminin, alpha 4) as a high-confidence effector gene underpinning a colocalizing genomic locus on chromosome 6 (PP4=0.89). The 95% credible set for the causal variant from the colocalization analysis consists of 36 variants (**Figure 3C**). All variants show opposing risk-increasing alleles for type 2 diabetes and schizophrenia. Variants in the 95% credible set for the causal variant from the colocalization are associated with trunk fat mass as well as depressive and maniac episodes in bipolar disorder ^87,88^.

Laminins are a family of extracellular matrix glycoproteins that constitute basement membranes ^89^. They have been implicated in multiple biological processes including attachment, migration and organization of cells into tissues during embryonic development. *LAMA4* encodes the laminin subunit alpha-4 protein, a structural component that contributes to cell adhesion and tissue organization in various biological processes ^90^. Heterozygous mutations in *LAMA4* are causal to the syndromic disease *dilated cardiomyopathy-1JJ*. In addition, variants in this gene are associated with various phenotypes including BMI, insulin, bipolar disorder and myocardial infarction. LAMA4 is the target of ocriplasmin, a drug indicated for the treatment of symptomatic vitreomacular adhesion ^91^.

*Lama4* knockout mice show decreased body size and weight and abnormal neuromuscular synapse morphology and lethargy. In humans, decreased body weight and lethargy are associated with decreased risk of type 2 diabetes and increased risk of schizophrenia, respectively. *LAMA4* shows differential expression in pancreatic islets from healthy versus diabetes patients and in brains from schizophrenia patients versus controls ^67,68^. It is over-expressed in diabetes patients and down-regulated in schizophrenia patients. The observed direction of effect of *LAMA4* on each investigated condition in knockout mice and in differential gene expression is concordant.

### Insights into disease biology and treatment targets

We conducted pathway enrichment analyses on the set of likely effector genes (**Table S8**) and find “homeostatic process” and “intracellular lipid transport” to be the most significantly enriched biological processes. The likely effector genes exhibited enrichment in further pathways related to lipids, for instance “lipid transport”, “lipid localization” and “lipid homeostasis”, as well as pathways related to the regulation of metabolic processes (**Figure 4**). These findings point toward a potential involvement of lipid metabolism in the comorbidity of type 2 diabetes and schizophrenia.

**Figure 4:**
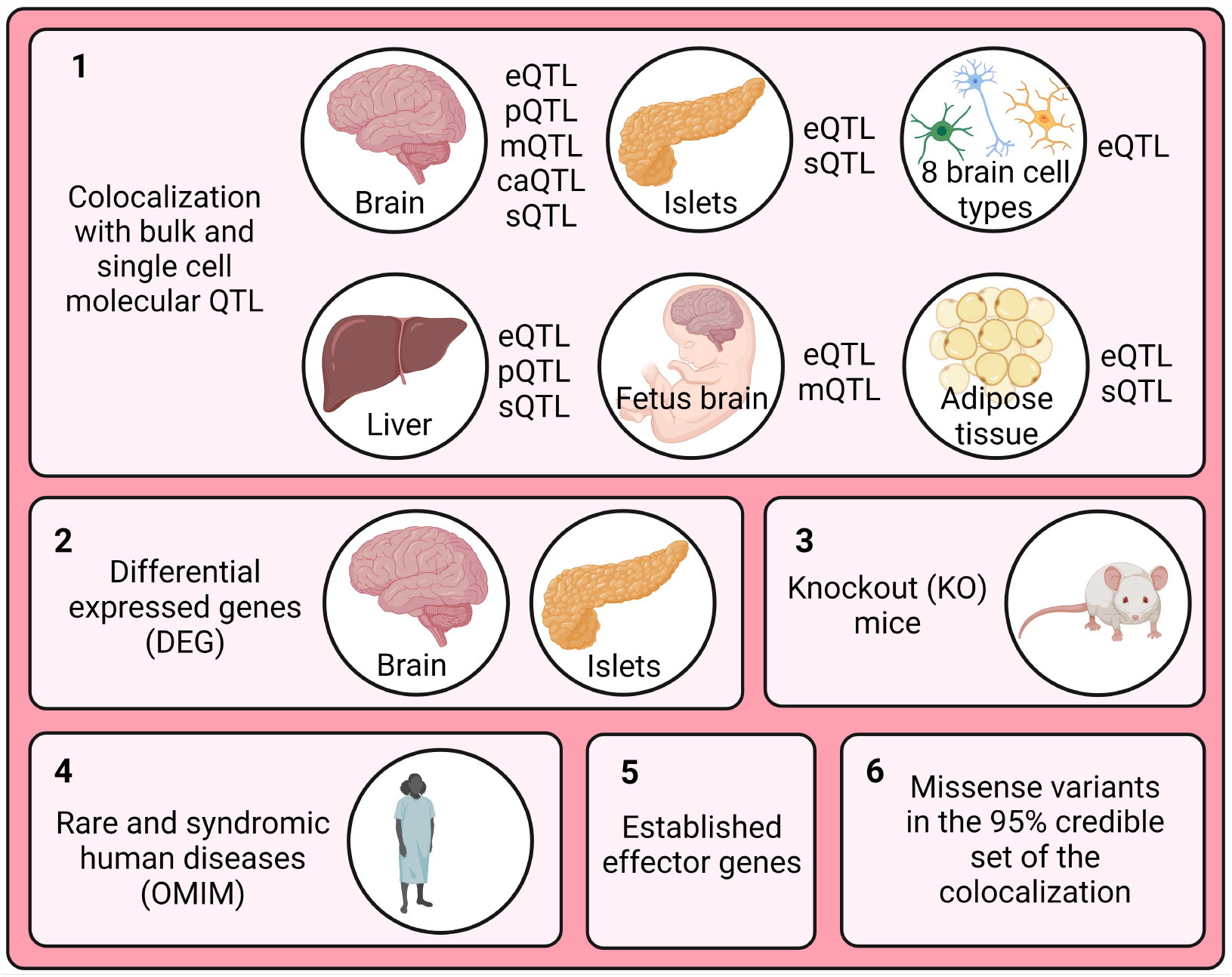
Results of enrichment analysis on the likely effector genes set.

The 15 shared likely effector genes for type 2 diabetes and schizophrenia represent relevant candidates for further functional and clinical investigation. Thus, we queried the druggability status of these genes and find that five are included in the druggable genome ^74^. Of these, four genes (*ATP1A1, GRIN2B, LAMA4* and *TK1*) are tier 1 druggable targets, i.e., their products are targets of existing drugs with market authorization or in clinical development, and one (*ACACA*) is a tier 2 druggable target, i.e., its product has known bioactive drug-like binding partners.

*ACACA* (acetyl-CoA carboxylase alpha) codes for the enzyme acetyl-CoA carboxylase alpha (ACC-alpha), which plays a role in fatty acid synthesis in humans, a process that impacts energy balance and lipid metabolism. In cases of impaired ACC-alpha function, biotin supplementation can help support proper enzyme activity as a cofactor, leading to better overall metabolic health ^92^. An ACC inhibitor was tested in a phase II clinical trial for type 2 diabetes and there is evidence that the ACC-alpha pathway in pancreatic alpha cells could be a therapeutic strategy in type 2 diabetes by limiting glucagon secretion ^93^.

*ATP1A1* (ATPase Na+/K+ transporting subunit alpha 1) encodes a subunit of the sodium-potassium pump, which plays a vital role in maintaining cell membrane potential and various physiological processes, including nerve impulse transmission ^94^. Cardiac glycosides drugs, such as digoxin, inhibit the sodium-potassium pump and have been used to treat heart-related conditions by enhancing the heart muscle contractions ^95^. Digoxin was also tested in a phase I clinical trial for epilepsy, depressive disorder and type 2 diabetes. In type 2 diabetes patients, it increased glucose intolerance ^96^. Digoxin augmented the effect of antiepileptic drugs in mice, increased the chances of depression after a myocardial infarction and was linked to increased risk of psychosis ^97-99^.

*GRIN2B* (glutamate ionotropic receptor N-methyl-D-aspartate type subunit 2B) encodes a subunit of the N-methyl-D-aspartate (NMDA) receptor ion channel in the brain that plays a role in synaptic plasticity, learning, memory, and various neurological functions ^100^. There are several approved drugs that modulate NMDA receptor activity, including felbamate and haloperidol. Felbamate, an anticonvulsant used to treat severe epilepsy, has an antagonistic effect on NMDA receptors, reducing their activity ^101^. Haloperidol is an antipsychotic used in the treatment of schizophrenia that has been shown to inhibit the NMDA receptor ^102^. Two further clinically approved NMDA antagonists, ketamine and acamprosate, have been tested for schizophrenia treatment with inconclusive results ^103,104^. Although *GRIN2B* is not rated as a high-confidence effector gene for schizophrenia, it has been shown that variants in this gene are associated with this condition in a Siberian and a Han Chinese population ^105,106^.

*LAMA4* is classified as a Tier 1 target in the druggable genome as the protein product of this gene is a target of the proteolytic enzyme ocriplasmin. Ocriplasmin is approved to treat symptomatic vitreomacular adhesion and was in clinical trial for stroke and diabetic macular edema treatment ^91^ (www.clinicaltrials.gov). LAMA1, LAMA3 and LAMA5, other laminins, are targets of the approved drug lanoteplase, used in the treatment of myocardial infarction but with unknown mechanisms of action ^107^. In an analysis aimed at identifying potential drug candidates based on perturbed transcriptomic pathways in Parkinson’s disease, lanoteplase emerged as the candidate with strongest enrichment ^108^.

## Discussion

In this work, we dissect the genetic etiology shared between type 2 diabetes and schizophrenia. In line with smaller-scale previous studies, we find evidence of a weak negative genetic correlation and no evidence of a causal relationship between the two comorbid conditions ^18,27^. We provide evidence of a bi-directional protective effect between BMI and schizophrenia. This association is in the opposite direction to the well-established causal effect between BMI and type 2 diabetes, pointing to potentially different adiposity-related mechanisms underpinning each condition. Since BMI exerts an effect on both conditions, then type 2 diabetes liability will influence schizophrenia and vice versa ^109^.

We replicated previous results showing that the causal effect of genetically predicted childhood BMI on type 2 diabetes is almost entirely attenuated when adjusting for adulthood BMI^77^. For schizophrenia, we observe the same trend, albeit with overall weaker effects. Schizophrenia is associated with low late-pregnancy maternal BMI, low birth weight and being thin during childhood^110^. Thus, the effect of body size on schizophrenia might be mediated earlier in development and is not fully captured by our analysis using genetically predicted data for early life BMI at 10 years old.

By leveraging recent large-scale GWAS for type 2 diabetes and schizophrenia, we find evidence of colocalization at 11 genomic loci. We score genes in the vicinity of colocalized regions by incorporating multi-omics and functional genomics information and derive a list of three high-confidence effector genes shared between both conditions. All three genes show opposite direction of effect on type 2 diabetes and schizophrenia risk. We show that the identified likely effector genes are enriched for biological pathways associated with lipid regulation. These results are in support of a causal effect of BMI on both conditions in opposite directions.

The biological function of the high-confidence genes *NUS1* and *LAMA4* as well as the result of the enrichment analysis highlight adipogenesis and cholesterol trafficking as potential biological mechanisms influencing the comorbidity between type 2 diabetes and schizophrenia. These mechanisms might reflect the presence of an underlying metabolic vulnerability in a subset of individuals with schizophrenia beyond the side effects of antipsychotics.

The genomic locus that yields *LAMA4* as a high-confidence effector gene does not reach nominal significance (p-value < 1 × 10^−^) in the multi-ancestry GWAS for type 2 diabetes (**Figure 3C**). The lead causal variant rs6568685 is an intron variant of *TRAF3IP2-AS1*. This variant reaches a p-value of 1.7 × 10^−^ in the European-ancestry type 2 diabetes GWAS and a p-value of 0.26 in the East Asian ancestry GWAS. Hence, this may be a European-specific association or a low frequency variant in East Asians. Alternatively, the association in this locus may reach genome-wide significance in diverse non-European ancestry datasets once sample sizes are large enough.

Human genetics evidence has been shown to support the majority of FDA-approved drugs in 2021^111^. We investigated the potential for drug development or repurposing for genes prioritized by our analyses, for the comorbidity between type 2 diabetes and schizophrenia. We highlight two interesting candidates for drug repurposing opportunities that could be investigated for schizophrenia treatment: lanoteplase, which targets laminins and is currently used in the treatment of myocardial infarction; and felbamate, an NMDA receptor inhibitor used to treat severe epilepsy.

Our analysis provides genetic evidence backing up schizophrenia treatment with haloperidol, an NMDA receptor inhibitor targeting *GRIN2B*^*102*^. However, production of NMDA receptor autoantibodies by the immune system can lead to NMDA receptor autoantibody-mediated psychosis, a type of autoimmune encephalitis ^112^. Additionally, studies have shown that functional NMDA receptors are also expressed in insulin producing cells^113^. Targeting NMDA receptors in human pancreatic islets with dextromethorphan, an NMDA receptor antagonist, has been found to lower blood glucose levels and increase insulin secretion^114^. The use of NMDA receptor antagonists to treat the comorbidity between type 2 diabetes and schizophrenia needs further investigation.

The majority of identified likely effector genes for the comorbidity between type 2 diabetes and schizophrenia have opposing direction of effect for the individual conditions. On the one hand, digoxin, a drug targeting *ATP1A1*, has been shown to increase glucose tolerance in type 2 diabetes patients ^96^, but on the other hand increase risk of psychosis ^99,115^. To avoid complications, drugs targeting genes with opposing direction of effect for different diseases should be administered with caution and alongside with monitoring of comorbidity risk and progression.

As a result of great efforts from the GWAS community, both the type 2 diabetes and schizophrenia GWAS summary statistics are meta-analyses that expand beyond European-centric data, by including data from diverse global populations. However, the vast majority of primary tissue molecular QTL data is derived from European-ancestry individuals. Hence, our analyses with statistical genetics methods that rely on LD estimations, such as Mendelian randomization, were constrained to using LD from European-ancestry populations, which still constitutes the majority (>70%) of samples contributing to GWAS. It will be important for future studies to generate molecular QTL data from a wide array of tissues across multiple diverse populations.

Mental health disorders are often accompanied by metabolic disorders that ultimately lead to premature death. Here we have studied the shared genetic underpinning between type 2 diabetes and schizophrenia and provide insights into the potential common biological mechanisms and shared effector genes. We have shown that the derived set of shared high-confidence effector genes have opposing direction of effect on the individual conditions. Our findings emphasize that treatments targeting those genes should be tempered with caution regarding exasperation of the comorbidity.

## Data Availability

All code produced in the present study are available upon reasonable request to the authors.

## Acknowledgments

Data on glycaemic traits have been contributed by MAGIC investigators and have been downloaded from www.magicinvestigators.org.

The Genotype-Tissue Expression (GTEx) Project was supported by the <u>Common Fund</u> of the Office of the Director of the National Institutes of Health, and by NCI, NHGRI, NHLBI, NIDA, NIMH, and NINDS.

G.M.K. and G.D.S. work in a unit funded by the Medical Research Council (MRC) and the University of Bristol (MC_UU_00032/01, MC_UU_00032/06).

G.M.K. acknowledges funding support from the Wellcome Trust (grant no: 201486/Z/16/Z and 201486/B/16/Z), the UK Medical Research Council (grant no: MC_UU_00032/06; MR/W014416/1; and MR/S037675/1), and the UK National Institute of Health Research Bristol Biomedical Research Centre (grant no: NIHR 203315).

LMH acknowledges funding from the National Institutes of Health: NIMH (R01MH124839, R01MH118278) and NIEHS (R01ES033630).

## Supplemental tables

**Table S1:** Genetic correlation analysis between type 2 diabetes (T2D) and schizophrenia (SCZ) using LD score regression.

**Table S2A:** Causal inference analysis between type 2 diabetes (T2D), schizophrenia (SCZ) and related traits using Mendelian randomization.

**Table S2B:** Steiger-filtered results of bi-directional Mendelian randomization analysis between type 2 diabetes (T2D), schizophrenia (SCZ) and related traits.

**Table S2C:** European-descent only sensitivity analysis of causal inference analysis between type 2 diabetes (T2D), schizophrenia (SCZ) and related traits using Mendelian randomization.

**Table S3A:** Causal inference analysis between childhood and adulthood body mass index (BMI) and type 2 diabetes (T2D) or schizophrenia (SCZ) using univariate Mendelian randomization.

**Table S3B:** European-descent only sensitivity analysis of causal inference analysis between childhood and adulthood body mass index (BMI) and type 2 diabetes (T2D) or schizophrenia (SCZ) using univariate Mendelian randomization.

**Table S4A:** Causal inference analysis between childhood and adulthood body mass index (BMI) and type 2 diabetes (T2D) or schizophrenia (SCZ) using multivariate Mendelian randomization.

**Table S4B:** European-descent only sensitivity analysis of causal inference analysis between childhood and adulthood body mass index (BMI) and type 2 diabetes (T2D) or schizophrenia (SCZ) using multivariate Mendelian randomization.

**Table S5:** Overview of genomic loci that colocalize between type 2 diabetes and schizophrenia with a posterior probability (PP4) > 0.8.

**Table S6:** Scoring of all genes within genomic loci that colocalize between type 2 diabetes (T2D) and schizophrenia (SCZ).

**Table S7A:** Causal inference analysis between expression of high confidence effector genes in disease-relevant tissues and type 2 diabetes (T2D) or schizophrenia (SCZ) using Mendelian randomization.

**Table S7B:** European-descent only sensitivity analysis of causal inference analysis between expression of high confidence effector genes in disease-relevant tissues and type 2 diabetes (T2D) or schizophrenia (SCZ) using Mendelian randomization.

**Table S8:** Gene set enrichment analysis on the likely effector genes using the Gene Ontology human networks via ConsensusPathDB.

**Table S9A:** List of knockout (KO) mice phenotypes and OMIM terms related to type 2 diabetes.

**Table S9B:** List of knockout (KO) mice phenotypes and OMIM terms related to schizophrenia.

